# Plasma proteomics for novel biomarker discovery in childhood tuberculosis

**DOI:** 10.1101/2024.12.05.24318340

**Authors:** Andrea Fossati, Peter Wambi, Devan Jaganath, Roger Calderon, Robert Castro, Alexander Mohapatra, Justin McKetney, Juaneta Luiz, Rutuja Nerurkar, Esin Nkereuwem, Molly F. Franke, Zaynab Mousavian, Jeffrey M. Collins, George B. Sigal, Mark R. Segal, Beate Kampman, Eric Wobudeya, Adithya Cattamanchi, Joel D. Ernst, Heather J. Zar, Danielle L. Swaney, the COMBO Study

## Abstract

Failure to rapidly diagnose tuberculosis disease (TB) and initiate treatment is a driving factor of TB as a leading cause of death in children. Current TB diagnostic assays have poor performance in children, and identifying novel non-sputum-based TB biomarkers to improve pediatric TB diagnosis is a global priority. We sought to develop a plasma biosignature for TB by probing the plasma proteome of 511 children stratified by TB diagnostic classification and HIV status from sites in four low- and middle-income countries, using high-throughput data-independent acquisition mass-spectrometry (DIA-PASEF-MS). We identified 47 proteins differentially regulated (BH adjusted p-values < 1%) between children with microbiologically confirmed TB and children with non-TB respiratory diseases (Unlikely TB). We further employed machine learning to derive three parsimonious biosignatures encompassing 4, 5, or 6 proteins that achieved AUCs of 0.86-0.88 all of which exceeded the minimum WHO target product profile accuracy thresholds for a TB screening test (70% specificity at 90% sensitivity, PPV 0.65-0.74, NPV 0.92-0.95). This work provides insights into the unique host response in pediatric TB disease, as well as a non-sputum biosignature that could reduce delays in TB diagnosis and improve detection and management of TB in children worldwide.

## Introduction

Tuberculosis (TB) is the leading cause of mortality from an infectious disease worldwide, with 10.6 million cases and 1.3 million deaths each year^1^. Children suffer a disproportionate burden: 12% of TB disease occurs in children, but children account for 16% of TB deaths. This disparity is largely due to delays in diagnosis and proper treatment initiation, as 96% of deaths are in children for whom treatment had not been initiated. While sputum-based diagnostic testing is routinely performed in adults, children are unable to reliably expectorate sputum, and sputum induction is typically required. Moreover, microbiological testing has sub-optimal sensitivity due to paucibacillary disease with low bacterial burden in children^2^. As a consequence, there is a large case detection gap where an estimated half of the children with TB disease, and two-thirds of those less than 5 years old, are not reported to public health programs. Consequently, the development of non-sputum biomarker TB tests is a global priority to improve TB diagnosis in children.

Host plasma protein biosignatures have shown promise for TB screening in adults, and have the potential to be translated into a simple point-of-care test^3–6^. However, these signatures have been largely derived from adult samples which poorly translate to pediatric TB disease due to different immune responses and disease manifestations in children. The development of pediatric-specific host biosignatures is a global priority for early detection of pediatric TB cases^7^. While mass spectrometry (MS) based proteomic analysis enables a broad untargeted approach to biomarker discovery, previous plasma proteomics efforts to identify plasma biosignatures of pediatric TB disease have been limited by small sample size, variable reference standards, and exclusive use of healthy controls that overestimate performance by selection of general inflammation markers rather than TB-specific markers^5,8^. Past studies also frequently utilized samples from a single region, leading to the discovery of candidate biomarkers that may reflect the co-morbidities and environment specific to the setting, and that fail to validate elsewhere. Here we have utilized high-throughput plasma proteomics and well-characterized pediatric TB cohorts across four countries to derive a host-based biomarker signature aimed at differentiating childhood TB disease from other causes of respiratory disease.

## Results

### Clinical Characteristics of the Cohort

We included plasma samples from 511 children with presumptive pulmonary TB from The Gambia (n=120), Peru (n=100), South Africa (n=111), and Uganda (n=180), of which 133 (26%) had microbiologically Confirmed TB, 120 (23.4%) Unconfirmed TB (clinically diagnosed), and 231 (45.2%) Unlikely TB (non-TB LRTI) based on NIH consensus definitions. In addition, we included 27 asymptomatic healthy children, of whom 8 (30%) had evidence of Latent TB infection based on a positive QuantiFERON-Gold test. Demographic and clinical characteristics are summarized in **Table 1** and provided for each patient in Supplementary Table 1; the median age was 4 years (IQR 2-7), 46.4% were female, 11.2% were living with HIV, and 52.6% were underweight.

**Table 1.** Cohort demographic and clinical characteristics (N=511)

|  | Age in years<br>(median, IQR) | <5 years<br>n (%) | 5-14 years<br>n (%) | Female<br>n (%) | HIV<br>infected<br>n (%) | Underweight<sup>#</sup><br>n (%) |
| --- | --- | --- | --- | --- | --- | --- |
| Confirmed TB (n=133, 26%) | 3 (1-7) | 77 (57.9%) | 56 (42.1%) | 64 (48.1%) | 23 (17.3%) | 83 (62.4%) |
| Unconfirmed TB (n=120, 23.4%) | 3 (2-7) | 82 (68.3%) | 38 (31.7%) | 53 (44.2%) | 16 (13.3%) | 69 (57.5%) |
| Unlikely TB (n=231, 45.2%) | 4 (2-8) | 129 (55.8%) | 102 (44.2%) | 109 (47.2%) | 18 (7.8%) | 117 (50.6%) |
| Healthy Control no TB infection<br>(n=19, 3.7%) | 5 (3.5-6.5) | 7 (36.8%) | 12 (63.2%) | 7 (36.8%) | 0 | 0 |
| Healthy Control with Latent TB<br>infection (n=8, 1.6%) | 4 (3-6.3) | 5 (62.5%) | 3 (37.5%) | 4 (50%) | 0 | 0 |
| All (N=511) | 4 (2-7) | 300 (58.7%) | 211 (41.3%) | 237 (46.4%) | 57 (11.2%) | 269 (52.6%) |

<sup>#</sup>Underweight defined as weight-for-age Z score < -2 if less than 5 years old, or body mass index < 18.5 if 5-14 years.

### DIA-PASEF enabled high-throughput and comprehensive plasma proteomics

For all children, we started from 1uL of undepleted plasma and performed high-throughput proteomics sample preparation using a filter-based processing in 96-well plates^10^, and followed by mass spectrometry analysis on a Bruker TimsTOF Pro mass spectrometer operating in data-independent acquisition mode (DIA-PASEF) (**Fig. 1A**)^11^. In total, we quantified 7,102 peptides and 859 proteins using a high throughput (30 min sample-to-sample) DIA-PASEF acquisition (**Fig. 1B**), with an average detection of 2,628 peptides and 498 proteins per sample (**Fig. 1C-D**, Supplementary Table 2). From this analysis, we removed 7 outlier samples showing low numbers of peptides and proteins, resulting in 504 samples in total. We achieved an average data completeness of 60.4%, with 241 proteins detected in all 504 samples and 411 detected in more than 75% of the samples (**Fig. 1E**). The concentration of proteins in plasma exists over a wide dynamic range exceeding 10 orders of magnitude, with a subset of proteins having very high concentrations (e.g. albumin) that can preclude the detection of lower abundance proteins. As we did not use immunodepletion to remove proteins of high concentration^12^, we evaluated the dynamic range in proteins detected in our proteomics experiments using reference concentration values from antibody and MS based-assays (HumanProteinAtlas^9^). Based on this analysis, we were able to quantify proteins over >4 orders of magnitude and down to a level of 12.1 ng/L concentration (SERPINF2), with a median concentration of 40 ng/L (**Fig. 1F**).

**Figure 1.**
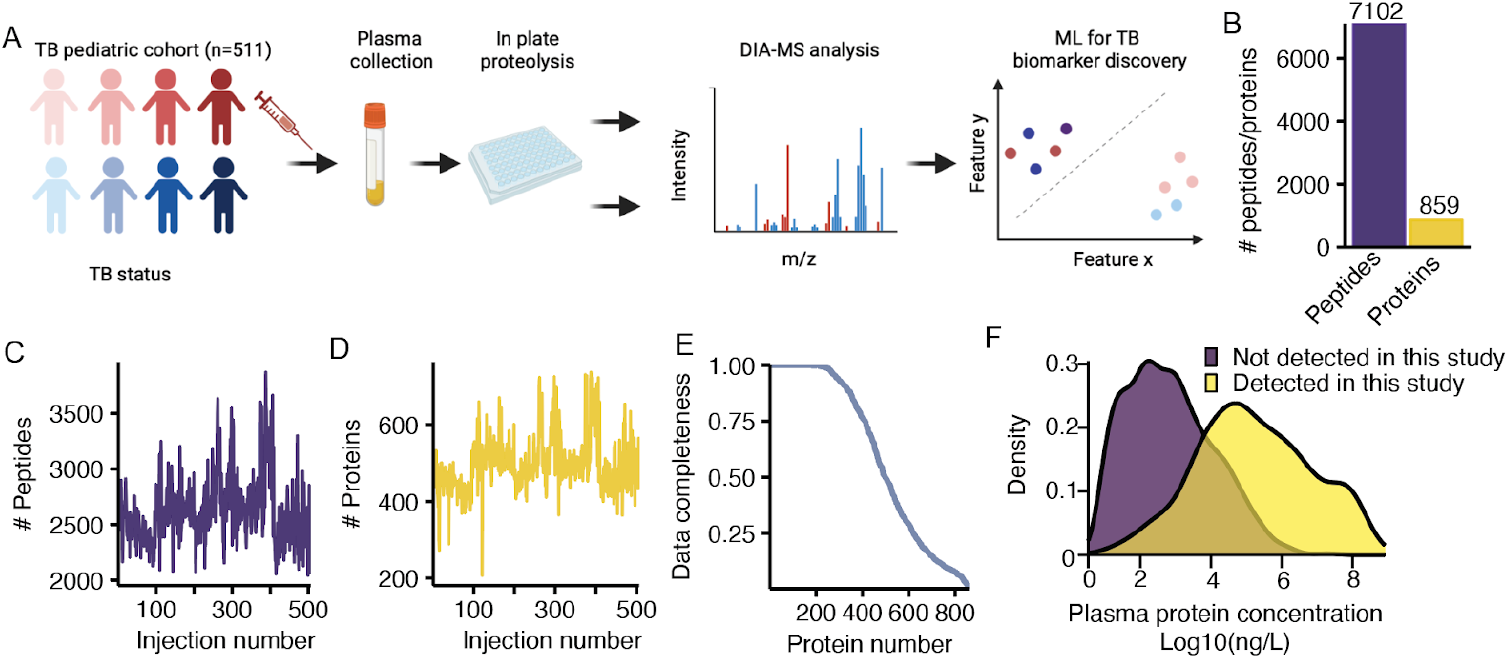
A high-throughput workflow for plasma proteomics. **A.** Plasma proteomics workflow and experimental design. **B**. Barplot showing the total number of unique peptide sequences and protein groups identified. **C-D**. Number of peptides (**C**), and proteins (**D**) identified per MS injection. **E**. Percentage of identifications (Y axis) versus the number of identified proteins (X axis). **F**. Density for the concentration range covered. X-axis represents the logged ng/L concentration determined from HumanProteinAtlas^9^, identified proteins are represented by the yellow density, while purple density represents remaining proteins.

We next evaluated our data across samples from the four clinical sites (**Fig. 2A**), and observed a consistent signal distribution of MS protein abundances, devoid of upper-end skewing, across 5 orders of magnitude (**Fig. 2B**). This resulted in highly consistent protein detections across countries, in which 88.7% of all proteins were detected across all sites, with less than 1% of all proteins displaying country-specific identification patterns (**Fig. 2C**). To normalize any variation between the various clinical sites, batches of sample preparation, or MS acquisition batches, we utilized COMBAT^13^, a parametric approach to mitigate batch effect commonly used in proteomics^14^. We used as batches the various clinical sites, with added covariates of the MS acquisition and sample preparation batches. After normalization and COMBAT correction, we reduced our data to two dimensions using single-value decomposition to visualize the sample distribution after PCA and positively reduced batch effects for most samples as exemplified by the majority of the samples not being separated by first or second component (**Fig. 2D**). Lastly, from a quantitative standpoint, we achieved a low coefficient of variation (CV) both within each country (average = 7.9%) and across all countries (∼8%) (**Fig. 2E**). Importantly, this analysis was performed using only proteins identified across more than 75% of the samples (n=411) to avoid artificially decreasing the CV due to the imputation process (see Methods). Overall, this suggests the absence of substantial country-specific protein abundance differences and the possibility of using the combined data from all clinical sites for analysis of TB specific differences.

**Figure 2.**
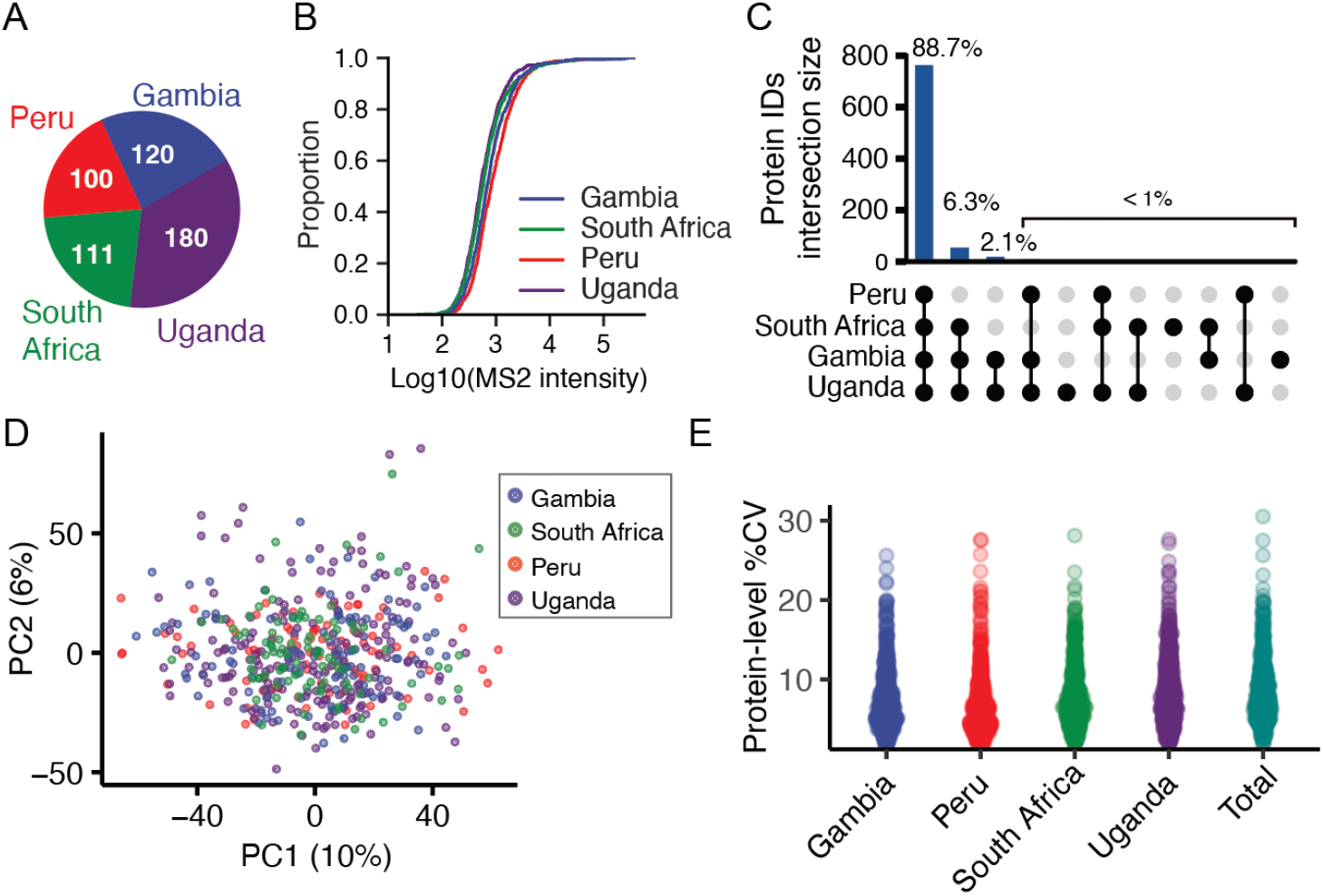
Quality control and reproducibility of plasma proteomics across multiple clinical sites. **A.** Pie chart illustrating the number of samples originating from each clinical site. **B**. Empirical cumulative distribution function plot for the raw MS intensity of the samples (X axis) from the various clinical sites. **C**. Upset plot showing the overlap in protein identifications between the different clinical sites. **D**. Principal Component Analysis (PCA) of the DIA-PASEF dataset following COMBAT batch correction. X axis shows the first component (10% variance) and Y axis the second component (6% variance). Each point represents a sample, while the color code indicates the clinical site. **E**. Protein level coefficient of variation within each clinical site and across all samples.

### Differential analysis to identify active TB candidate biomarkers

We first evaluated the ability of plasma proteomics to separate healthy children from symptomatic children undergoing evaluation for pulmonary TB by comparing the protein levels of known inflammatory markers. As expected, serum amyloid protein 1, 2, 4 (SAA1, SAA2, SAA4) and C-reactive protein (CRP) were all significantly upregulated among symptomatic children, with SAA2 displaying the greatest difference amongst the acute phase proteins (**Fig. 3A**). However these inflammatory markers were not able to significantly distinguish between the different groups of symptomatic children (i.e. Confirmed, Unconfirmed, or Unlikely TB) (**Fig. 3A**).

**Figure 3.**
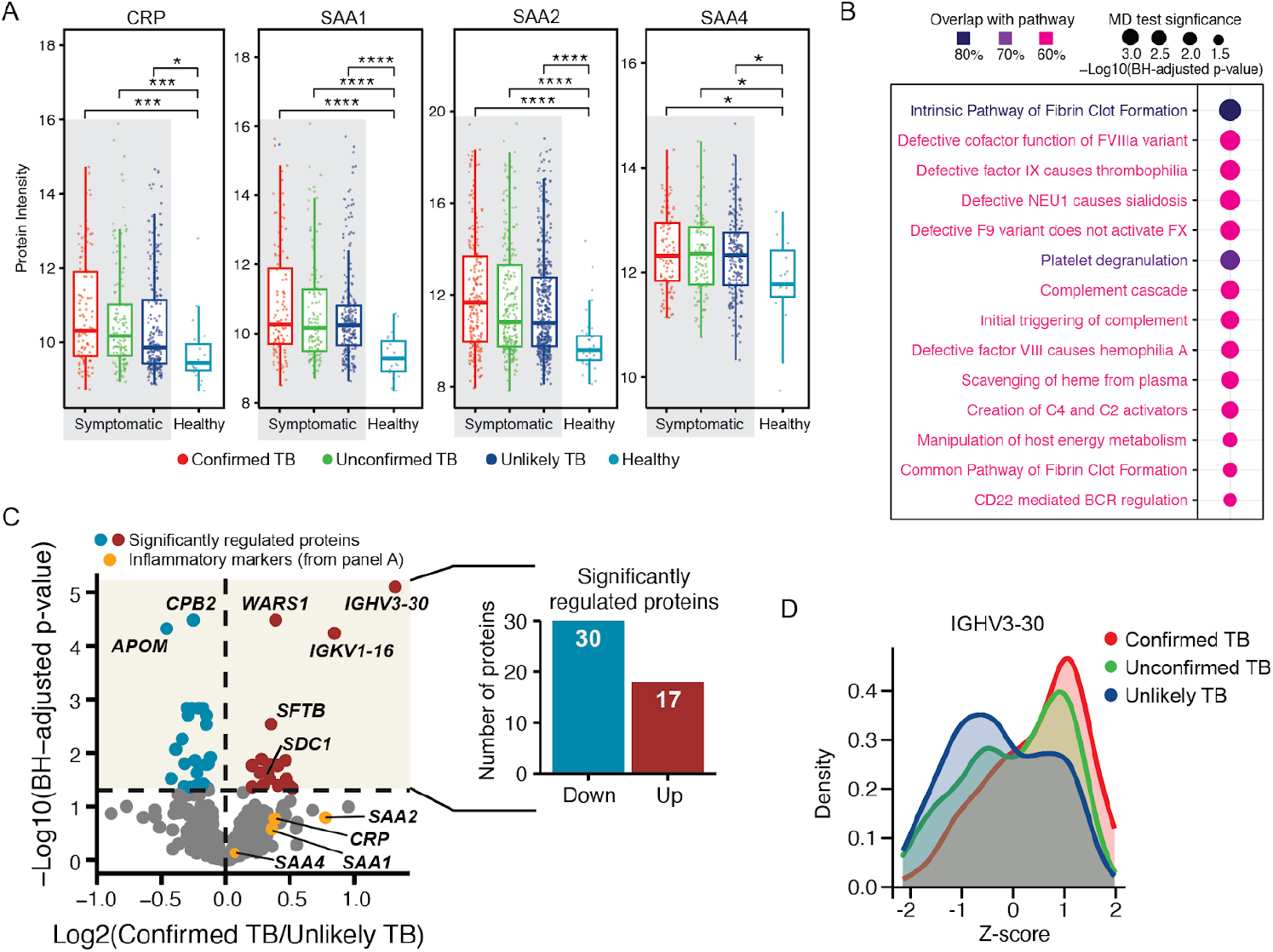
Abundance proteomics analysis of pediatric TB cohorts. **A.** Benchmark of data between patients with respiratory burden and healthy controls (excluding Latent TB Infection). X axis shows the TB classification status while Y axis represents the protein-level intensity. Box shows the IQR and its Kruskal-Wallis test is represented as ^*^for p < 0.01, ^***^ for p < 0.0001, and ^****^ for p < 0.000001. **B**. Gene set enrichment analysis for identification of dysregulated pathways between Confirmed TB and Unlikely TB. Dot size represents the Benjamini-Hochberg (BH) adjusted p from a mean difference (MD) test. Colors indicate the overlap between each signaling pathway and the protein dataset. **C**. Volcano plot between Confirmed and Unlikely TB. The X-axis shows the Log2 fold change at the protein level, while the Y-axis represents the significance as −log10 of the BH corrected p-values. Significant proteins (BH-adjusted *p* < 5%) are shown in red and blue. Barplot showing the number of differentially expressed proteins (DEPs) divided in upregulated (red) and downregulated (blue). **D**. Density plot showing the z-scored intensity for the most significantly regulated protein (IGHV3-30), divided by TB status in confirmed TB (red), unconfirmed TB (green) and unlikely TB (blue).

We next focused on comparing plasma protein levels in children with Confirmed TB (n=112) and Unlikely TB (n=235) to identify biomarkers that could distinguish TB disease from other non-TB respiratory diseases. To identify pathways with dysregulated patterns between Confirmed TB and Unlikely TB, we performed a pathway enrichment analysis on each pathway included in the KEGG and REACTOME databases, using only gene sets with more than 50% of overlap with our plasma proteomic datasets. In total, 14 pathways showed significant differential means with Benjamini-Hochberg adjusted p-value < 0.05 (**Fig. 3B**, only pathways with over 60% overlap are represented). Amongst the pathways showing significant regulation, we identified several related to complement activation, which have also been identified in studies of whole blood transcriptomics in TB^15,16^. Complement upregulation in the context of TB may reflect activation of the classical pathway by antigen-antibody complexes, activation of the alternative pathway or mannose-binding lectin pathway by components of the Mtb cell envelope or cell wall, and/or through increased synthesis as acute phase proteins.

From this comparison between Confirmed and Unlikely TB, we identified 47 proteins displaying significantly different abundances, of which 30 displayed downregulation and 17 displayed upregulation (**Fig. 3C**, Supplementary Table 3). Interestingly, one of the the proteins displaying the most statistically significant regulation was the tryptophanyl t-RNA synthetase WARS1, which was increased in children with Confirmed TB vs. Unlikely TB (log2FC = 0.39, BH *adjusted p* = 3.3^*^10-5) (**Fig. 3C**), and is linked to TB infection via multiple mechanisms. Overall, the majority of these are known plasma proteins with previous classifications as secreted or extracellular proteins (38/48), minimizing the possibility of random variation in tissue leakage driving the distinction between groups. For the remaining 10 (WARS1, DBH, TUBA1A, ICAM1, GSN, LTA4H, SDC1, CSF1R, THBS4, CDH13), manual curation of their localization demonstrated the majority being potentially secreted (9/10) with only one (TUB1A1) not having reported extracellular localization.

We further identified upregulation of multiple specific immunoglobulin heavy (IGHV1-18, IGHV1-3, IGHV2-26, IGHV3-23, IGHV3-30) and light chain variable domains (IGKV1-16, IGKV1D-33 and IGKV3-20) across several countries (**Fig. 3D**, Supplementary Fig. S1) potentially suggesting an oligoclonal humoral response to TB disease. Additionally, we observed significantly different levels of several proteins (APOM, PON1, CPB2) (**Fig. 3C**), which have been previously identified in plasma proteomic studies of severe vs non severe COVID-19^17^ and an adult TB study^5^, potentially pointing towards those proteins as general markers of lung inflammation rather than specific markers of pediatric TB disease.

### Machine learning for identification of a host-protein biosignature of active pulmonary TB disease

To identify the smallest subset of features achieving the required target product profile (TPP) for a screening test (70% specificity at 90% sensitivity), we first utilized LASSO to reduce the number of features to a subset that would allow exhaustive brute-force approaches. The choice of LASSO over other feature selection approaches like Tree-based or recursive feature elimination (forward or reverse) was due to the inherent sparsity of the resulting solutions and the computational performance. We utilized LASSO using 20-fold cross-validation, which led to removal of a large portion of features resulting in 67 with non-0 LASSO coefficients (**Fig. 4A** and Supplementary Table S4). Notably, simply selecting the top N most important proteins by their LASSO feature importance did not reach the WHO TPP for any of the N utilized (Supplementary Fig. S2), which suggests the use of deep combinatorial analysis to evaluate the performance of a small subset of features.

**Figure 4.**
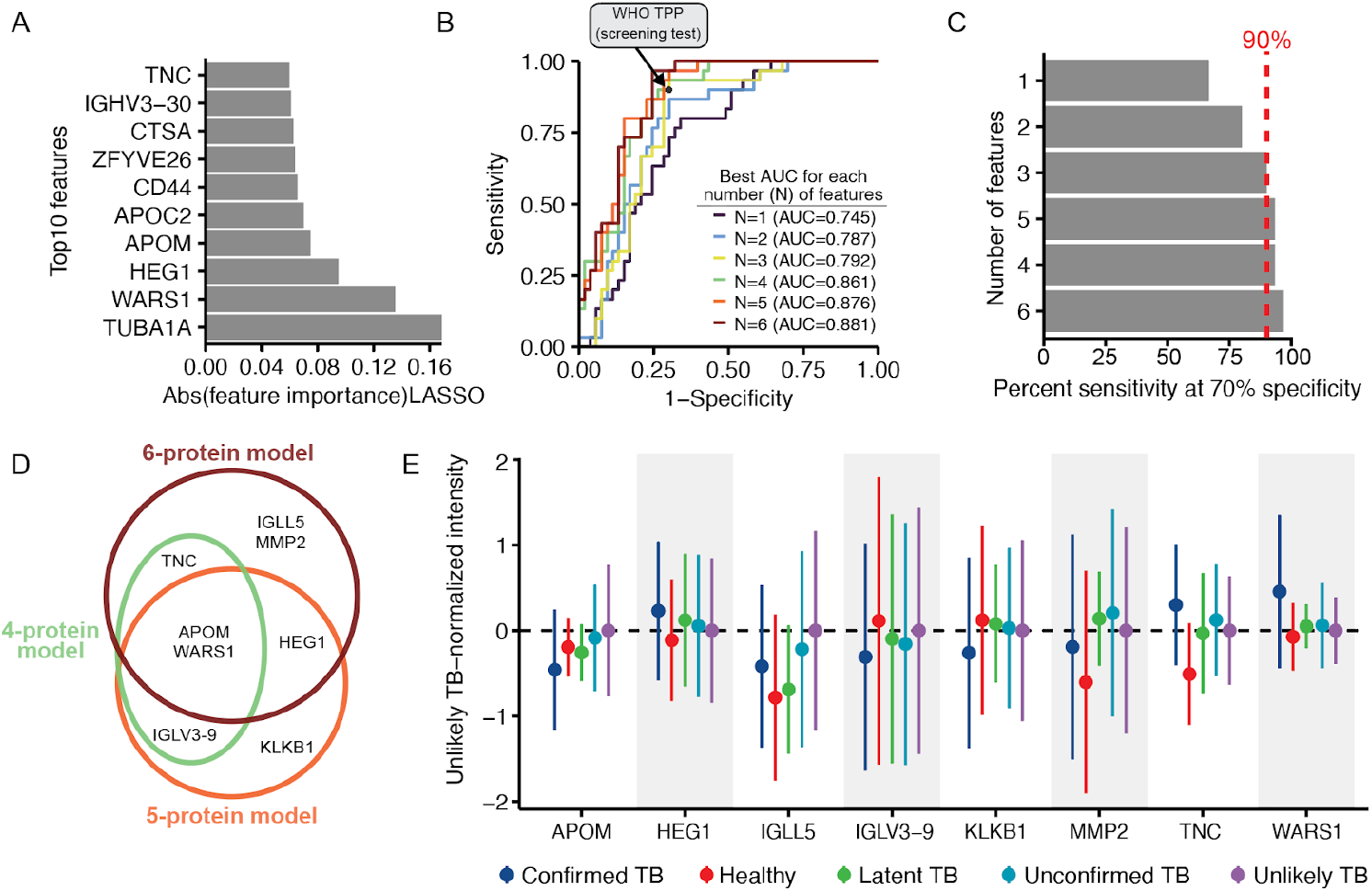
Machine learning to develop a parsimonious biosignature for pediatric TB disease. **A.** Absolute feature importance from a LASSO model for the top10 most important features. **B**. ROC curves for best-scoring combination of features. Each curve represents the feature subset achieving the highest AUC derived from all combinations of 1 (n=67), 2 (n=2210), 3 (n=47904,) 4(n=766479), 5 (n= 9657647) and 6 (n=99795695) features. WHO TPP for a screening test (70% specificity and 90% sensitivity) is denoted by the black circle. **C**. Barplot for the sensitivity achieved at 70% specificity for all 6 models. Dotted red line represents 90% sensitivity. **D**. Venn diagram of the overlap in proteins from the 4-, 5-, or 6-protein model. **E**. Dotplot representing the mean (dot) and the standard deviation (line) for the proposed biosignature proteins across the three models achieving the WHO TPP. Different colors highlight the different TB classes according to NIH consensus definition. Each protein is normalized to the Unlikely TB protein abundance for that respective protein.

Thus, we decided to investigate the best combination of a small subset of features using the WHO TPP as an objective function. Specifically, we calculated all possible combinations of N features (from 1 to 6) and selected the combinations maximizing the specificity at 90% sensitivity. We derived six linear models (trained on a 75% balanced subset of the data and tested on 25% of the remaining samples), of which three exceeded the WHO TPP for a screening test (**Fig. 4B**) consisting of 4, 5, or 6 proteins. The 4 and 5 proteins models achieved 93% sensitivity at 70% specificity (95% CI for 4 protein model 0.73-0.97, 95% CI for 5 protein model 0.78-0.99), and the 6 protein models achieved 96.7% sensitivity at 70% specificity (95% CI 0.83-0.99) on our test data (**Fig. 4C**, n=83, 30 positive, 53 negative). The derived features for the 4 to 6 protein models were mostly shared, with APOM and WARS1 being shared across the 4, 5, and 6 protein models (**Fig. 4D**). The selected proteins for most models showed small variance and significantly different means across all TB classes (**Fig. 4E**), potentially suggesting their relevance in TB disease. Two proteins further showed regulation when comparing Confirmed TB and Unlikely TB: WARS1 (log2FC 0.38, q=10x^-^5) and APOM (log2FC −0.45, q=10x^-^5) (**Fig. 4E**). Each individual showed a low AUC ranging from 0.577 (HEG1) to 0.745 (APOM), suggesting the lack of a single indicative feature driving the AUC and the need for at least 4 proteins to achieve the WHO TPP (Supplementary Fig. S3).

### Detection of Unconfirmed TB

We tested the derived biosignatures on 115 Unconfirmed TB cases that passed our proteomics quality control filtering to assess if we could further identify TB cases in symptomatic children with culture-negative disease. In this comparison we only used biosignatures exceeding the WHO TPP for a screening test (4, 5, and 6 protein models) and utilized as probability threshold for classification the AUC point that achieved the WHO TPP. The various models supported the diagnosis of TB in Unconfirmed TB (negative by sputum-based testing) in ∼30% of the cases, with the 4 protein model predicting 49/115 positive TB cases, the 5 protein model 35/115 cases and the 6 protein model 42/115 positives (**Fig. 5A**). We observed good agreement between predictions, with 26/115 samples (22%) positively predicted by all models and 42 (36%) positively predicted by at least 2 models (**Fig. 5B**). Importantly, we did not observe separation between healthy and latent TB when utilizing any of these three biosignatures, suggesting that these are specific for active TB disease (Supplementary Fig. S4). When evaluating the separation between the various Unconfirmed TB samples and the Confirmed TB group using all identified proteins, we observed a trend where samples positively predicted by the three models (n=26), clustered more closely to the Confirmed TB group in latent space derived by PCA, and showed separation on the first component from the negatively predicted unconfirmed samples (n=60) (**Fig. 5C**). This suggests that we robustly extrapolated a valid biosignature as the individual contribution of these 8 proteins on the total number of proteins identified (850) is small with only TNC ranking amongst the top 20% features driving the separation on the first component (Supplementary Fig. 5).

**Figure 5.**
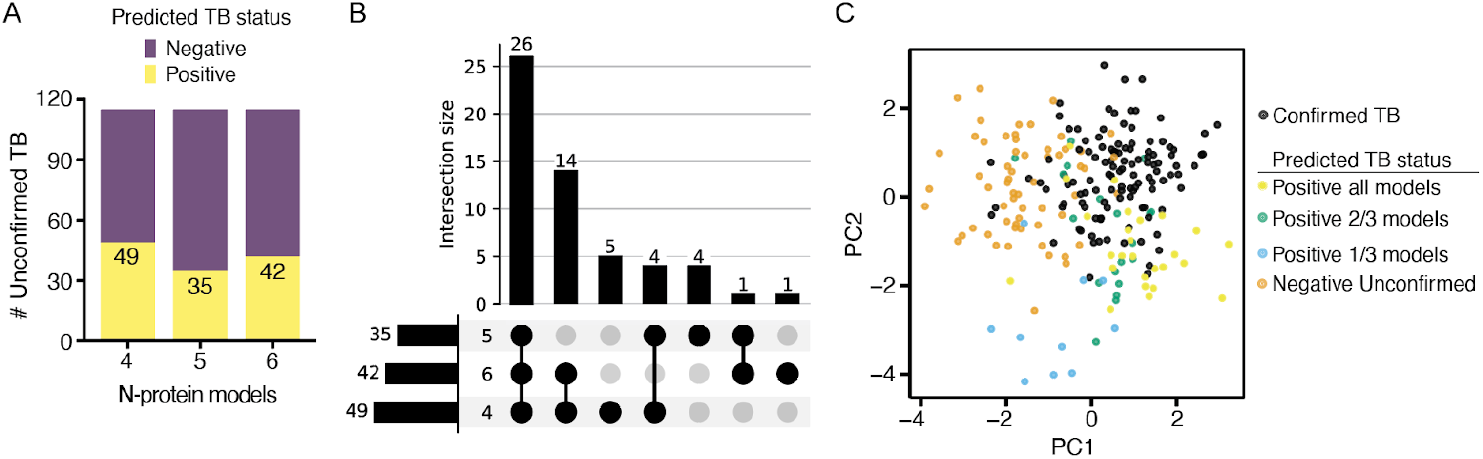
Detection of Unconfirmed TB. **A.** Barplot showing the number of positive predicted (yellow) and negative predicted (purple) in the proposed linear models using 4, 5, or 6 proteins. Values in the barplot indicate the number of positive predicted cases. **B**. Upset plot displaying the overlap between all predictions. **C**. Principal component analysis of Confirmed and Unconfirmed TB. X-axis shows the first component and Y-axis shows the second component. Each dot represents a sample. Samples are color coded based on either TB status (Confirmed TB, black) and further for the Unconfirmed TB based on the prediction of the various models (⅓ models cyan, ⅔ models green, or all models yellow). Samples negatively predicted by all models (Negative Unconfirmed) are shown in orange.

## Discussion

This study represents the largest TB plasma proteomics study in children to date, and encompasses a diverse pediatric cohort of >500 samples across clinical sites in four LMIC and two continents. The scale of this analysis was made possible by the use of data-independent acquisition to provide high-throughput, accurate, and precise quantification of hundreds of proteins within only ∼30 minutes of MS acquisition. This is in contrast to previous work for the development of host-based biomarker for TB using plasma proteomics, which have revolved around the use of proteomics multiplexing for quantification (e.g. ITRAQ) and long acquisition times, both of which are detrimental for acquisition of large clinical cohorts. Furthermore, the power of this study is amplified by our cohort design, which includes both healthy controls and >200 controls with non-TB respiratory diseases. The inclusion of a non-TB respiratory disease control group addresses a key clinical diagnostic challenge to distinguish children with pulmonary TB disease from those with symptoms due to other causes. Inclusion of this control group avoids the detection of candidate TB biomarkers that are non-specific inflammatory markers that cannot differentiate among symptomatic states, as observed with CRP, SAA1, SAA2, SAA3, and SAA4, and which were included in previous plasma proteomic biosignatures^5,8^.

An important milestone of this work is the application of machine learning to develop a minimal host-based biosignature consisting of 4-6 proteins that separate children with Confirmed TB vs. Unlikely TB at a level of specificity and sensitivity that meets or exceeds the WHO criteria for a TB screening test^19^. We found WARS1 to be a common protein among all protein biosignatures. WARS1 (also known as TrpRS) has previously been linked to TB infection by multiple mechanisms. First, upon *M. tuberculosis* infection, a multitude of lymphocytes, including CD4 and CD8 T cells, noncanonical T cells, natural killer cells, and type 1 innate lymphoid cells^19^ up-regulate interferon gamma (IFNγ) as part of the host immune response, which in turn induces WARS1 expression. WARS1 is also induced by tryptophan depletion. Tryptophan depletion by the kynurenine pathway has been detected in multiple metabolomic studies in active TB disease^20–23^, hence our data further supports previous reports on the importance of Tryptophan metabolism in active TB diseases versus other respiratory illnesses.

Importantly, our protein biosignatures did not separate between healthy children and children with latent TB infection for any of the tested models, suggesting that these protein biosignatures are specific for active TB disease. Moreover, application of these host-biosignatures to children with Unconfirmed TB was able to further support the diagnosis of TB in ∼30% of cases that were negative by sputum-based testing. While this work represents the first untargeted discovery-proteomics biosignature for childhood TB, there have been several cytokine-based signatures identified for TB in children. However, these targeted analyses were completed at a single center with a small sample size. For example, prior work identified a 3-cytokine signature to distinguish children with TB disease from other respiratory diseases in the Gambia, but they achieved a lower AUC of 0.74 and 72.2% sensitivity^18^. Our study benefited from a large sample size, representation from four countries, with a high proportion who were under five years old, living with HIV, and were undernourished. While further prospective validation and subgroup analyses are needed to evaluate robustness and reproducibility, our findings suggest that a simple host-based proteomic signature could be integrated into a point-of-care device for non-sputum TB screening for children.

In conclusion, untargeted proteomics was able to broadly evaluate the plasma of children across four countries, and identify candidate host protein biomarkers that could distinguish pediatric TB disease from other respiratory diseases. Moreover, from these candidate markers, we identified novel plasma protein biosignatures of only 4-6 proteins for childhood TB disease that achieved the minimum accuracy for a TB screening tool. These efforts have provided greater characterization of the unique immune response in pediatric TB disease, while providing a new non-sputum biosignature that could reduce delays in TB diagnosis and improve detection and management of TB in children worldwide.

## Data Availability

All data produced in the present work are contained in the manuscript or available online at the MassIVE repository with Dataset Identifier: MSV000096394.

https://massive.ucsd.edu/ProteoSAFe/static/massive.jsp

## Data availability

The supporting MS data is available via the MassIVE repository with Dataset Identifier: MSV00000096394, Reviewer Username: MSV000096394 _reviewer, and Reviewer Password: H6DVzA8T79W1RC6c.

## Acknowledgments

Figure 1A was created in BioRender by AF (2025) https://BioRender.com/o99n413. NIH R01AI152161 and R01AI175312 to AC, JDE, and DLS. NIH K23HL153581 to DJ. UKRI MR/P024270/1 and MR/K011944/1 to BK.

## Additional COMBO consortium contributors

Cynthia Baard, University of Cape Town, South Africa

Yekiwe Hlombe, University of Cape Town, South Africa

Lesley Workman, University of Cape Town, South Africa

Margaretha Prins, University of Cape Town, South Africa

Abdoulie Tunkara, The Gambia at the London School of Hygiene and Tropical Medicine, The Gambia

Binta Saidy, The Gambia at the London School of Hygiene and Tropical Medicine, The Gambia

Francis S. Mendy, The Gambia at the London School of Hygiene and Tropical Medicine, The Gambia

Madikoi Danso, The Gambia at the London School of Hygiene and Tropical Medicine, The Gambia

Marie P. Gomez, The Gambia at the London School of Hygiene and Tropical Medicine, The Gambia

Martina Boakarie, The Gambia at the London School of Hygiene and Tropical Medicine, The Gambia

Sarjo Koita, The Gambia at the London School of Hygiene and Tropical Medicine, The Gambia

Sheriff Kandeh, The Gambia at the London School of Hygiene and Tropical Medicine, The Gambia

Jascent Nakafero, World Alliance for Lung and Intensive Care Medicine in Uganda (WALIMU), Uganda

Gertrude Nanyonga, World Alliance for Lung and Intensive Care Medicine in Uganda (WALIMU), Uganda

Juliet Namboowa, World Alliance for Lung and Intensive Care Medicine in Uganda (WALIMU), Uganda

Winnie Nabakka, World Alliance for Lung and Intensive Care Medicine in Uganda (WALIMU), Uganda

Nsereko Moses, World Alliance for Lung and Intensive Care Medicine in Uganda (WALIMU), Uganda

Ainebyona Aggrey, World Alliance for Lung and Intensive Care Medicine in Uganda (WALIMU), Uganda

Alfred Andama, World Alliance for Lung and Intensive Care Medicine in Uganda (WALIMU), Uganda

Moorine sekadde, World Alliance for Lung and Intensive Care Medicine in Uganda (WALIMU), Uganda

Mary Mudiope, World Alliance for Lung and Intensive Care Medicine in Uganda (WALIMU), Uganda

Ezekiel Mupere, World Alliance for Lung and Intensive Care Medicine in Uganda (WALIMU), Uganda

Hellen Aanyu Tukamuhebwa, World Alliance for Lung and Intensive Care Medicine in Uganda (WALIMU), Uganda

## Author contributions

Conceptualization: DLS, DJ, AC, JDE. Data curation: AF, JM, PW, RC, JL, EN, MF, BK, EW, AC, DJ, HZ. Formal analysis: AF, JM, RC, RN, ZM. Funding acquisition: DLS, JDE, AC, DJ. Investigation: AF, PW, RC, JL, EN, MF, BK, EW, AC, JDE, DJ, HZ, AM. Methodology: AF, DJ, DLS, JDE, AC, JC, GBS, MRS. Project administration: DLS, AC, JDE, BK, EW, HZ. Software: AF. Resources: JDE, AC, DJ, HZ, MF, RC, BK, EW. Supervision: DLS, JDE, DJ, AC, MRS, BK, EW, HZ. Validation: AF, JM. Visualization: AF, JM, DLS, RN, ZM. Writing – original draft: AF, DLS, JDE, AC, DJ, RN, ZM. Writing – review & editing: All authors

## Supplementary figures

**Supplementary Figure 1.**
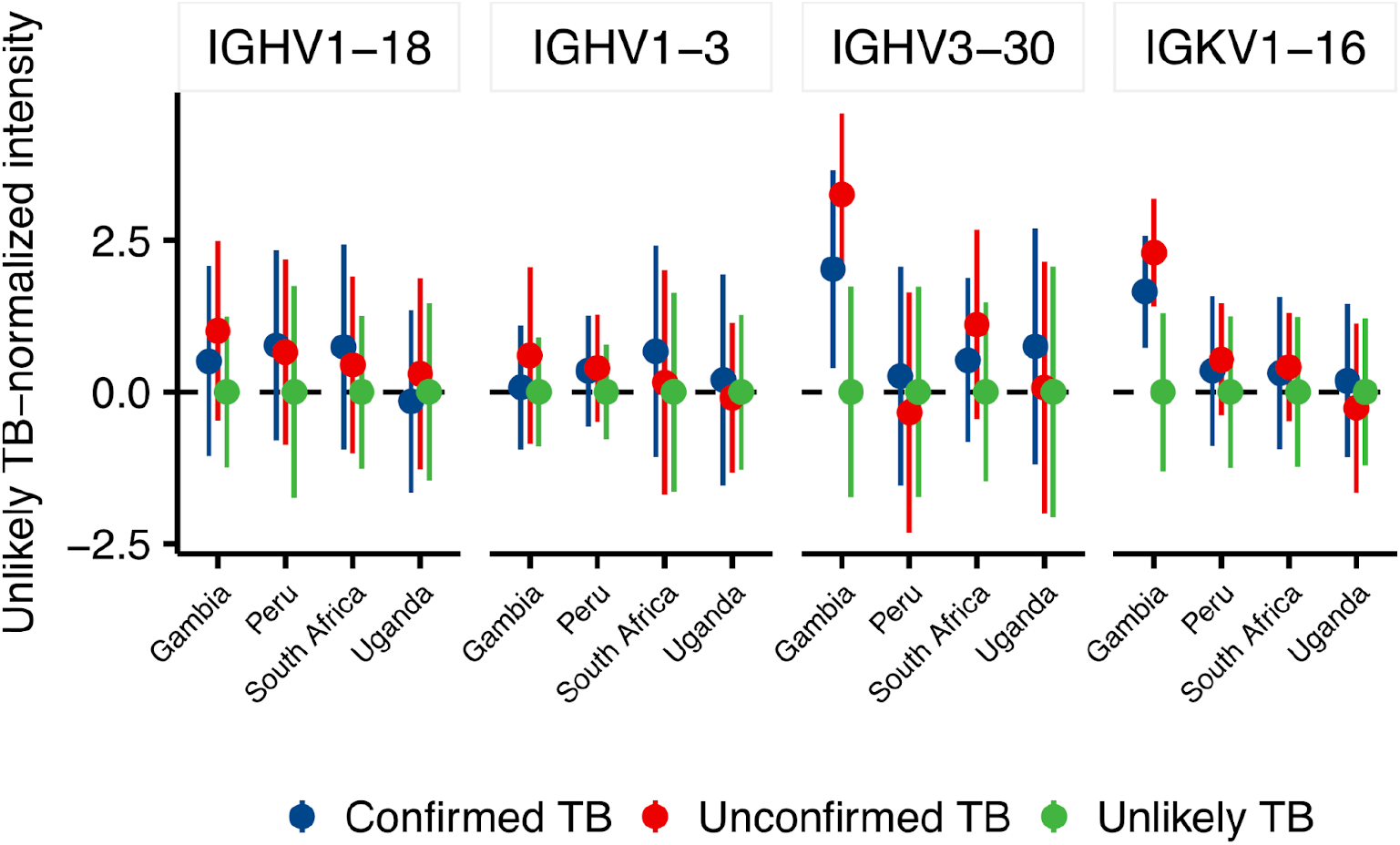
Upregulated IGg level, expressed as protein intensity normalized to the global amount of the Unlikely TB condition (Y axis) stratified by clinical site (X axis).

**Supplementary Figure 2.**
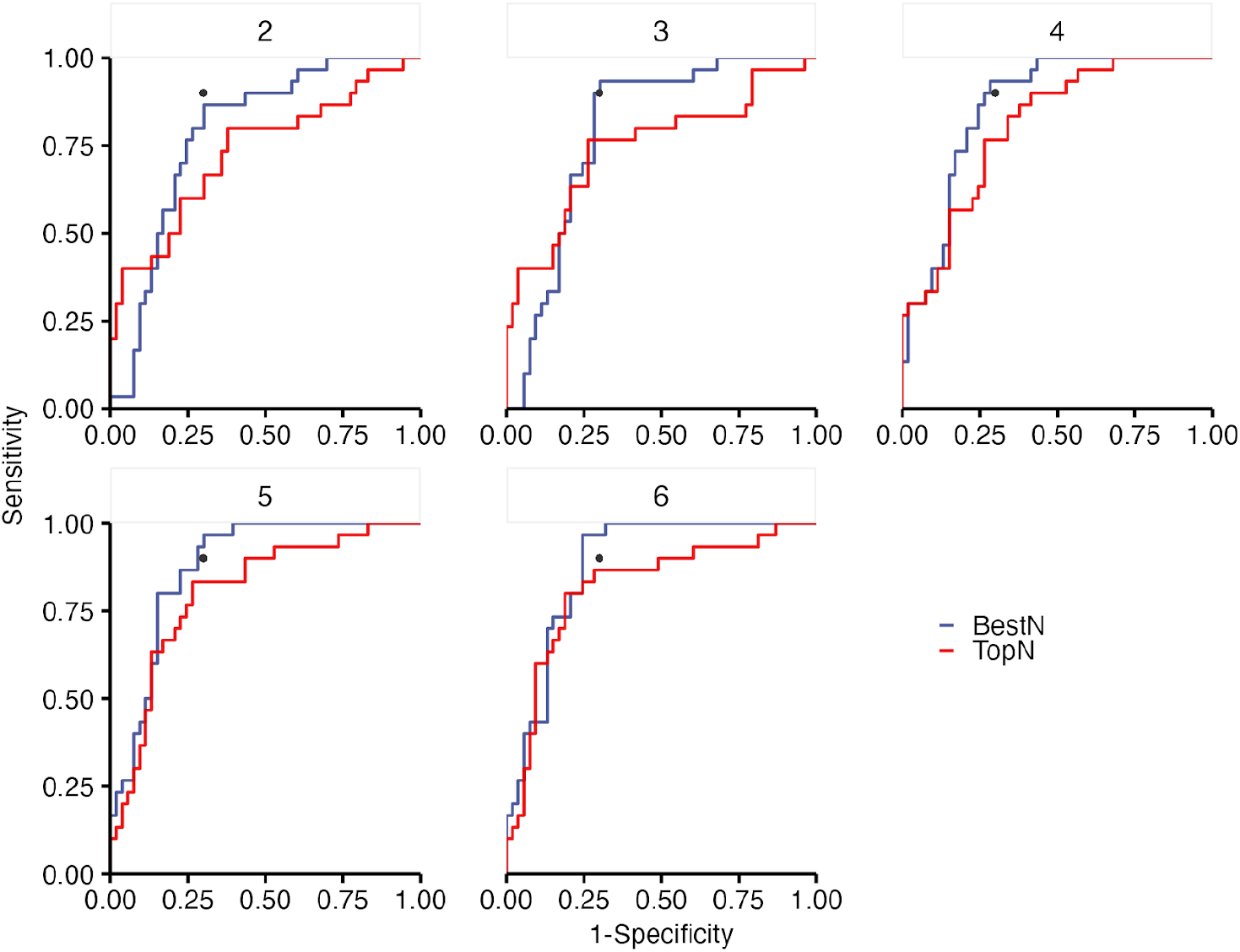
ROC Curves for various number of features comparing the best combination (BestN, blue line) to the corresponding number of most important ones from the LASSO feature importance (TopN, red line).

**Supplementary Figure 3.**
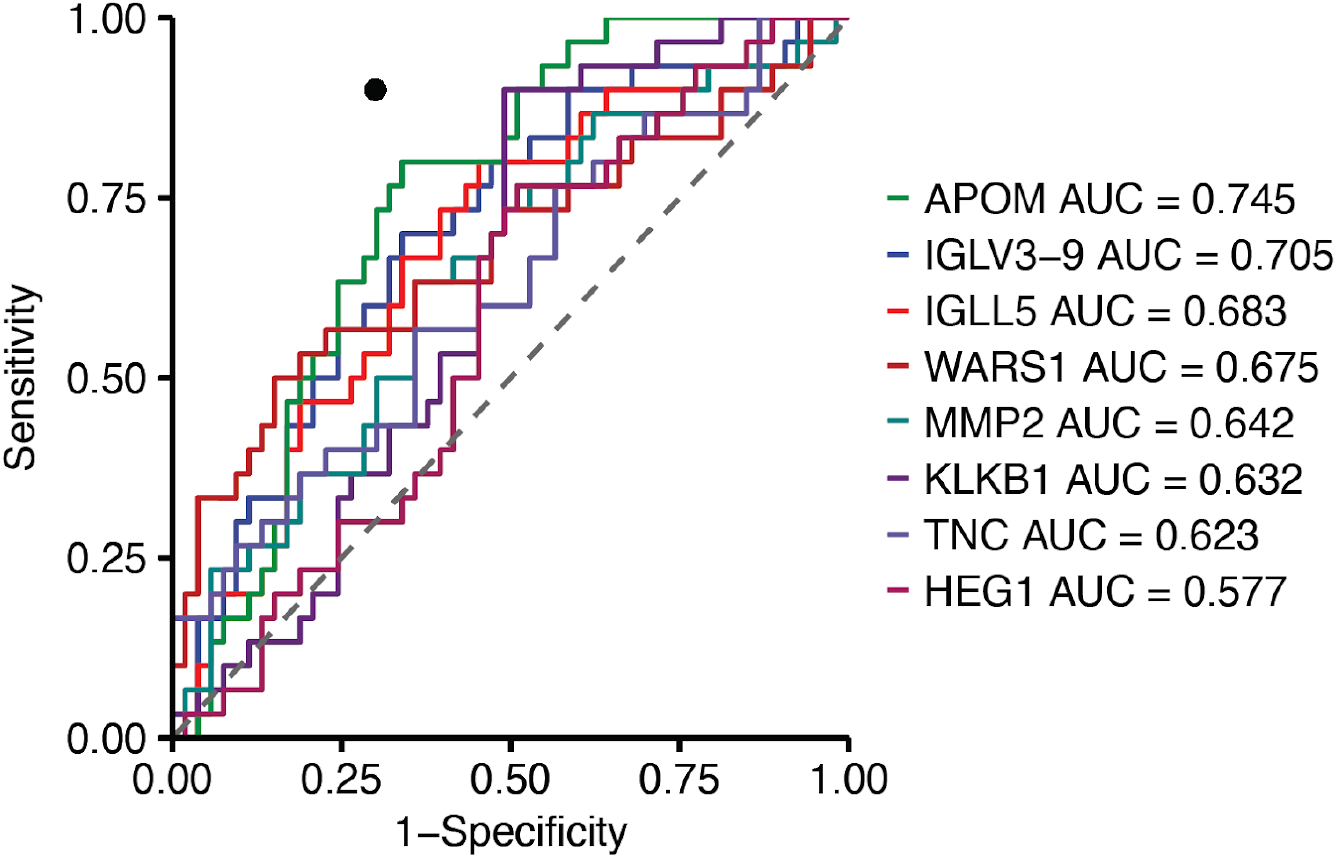
Individual ROC curves for the proteins from the developed biosignatures. The black circle designates the WHO target product profile sensitivity and specificity.

**Supplementary Figure 4.**
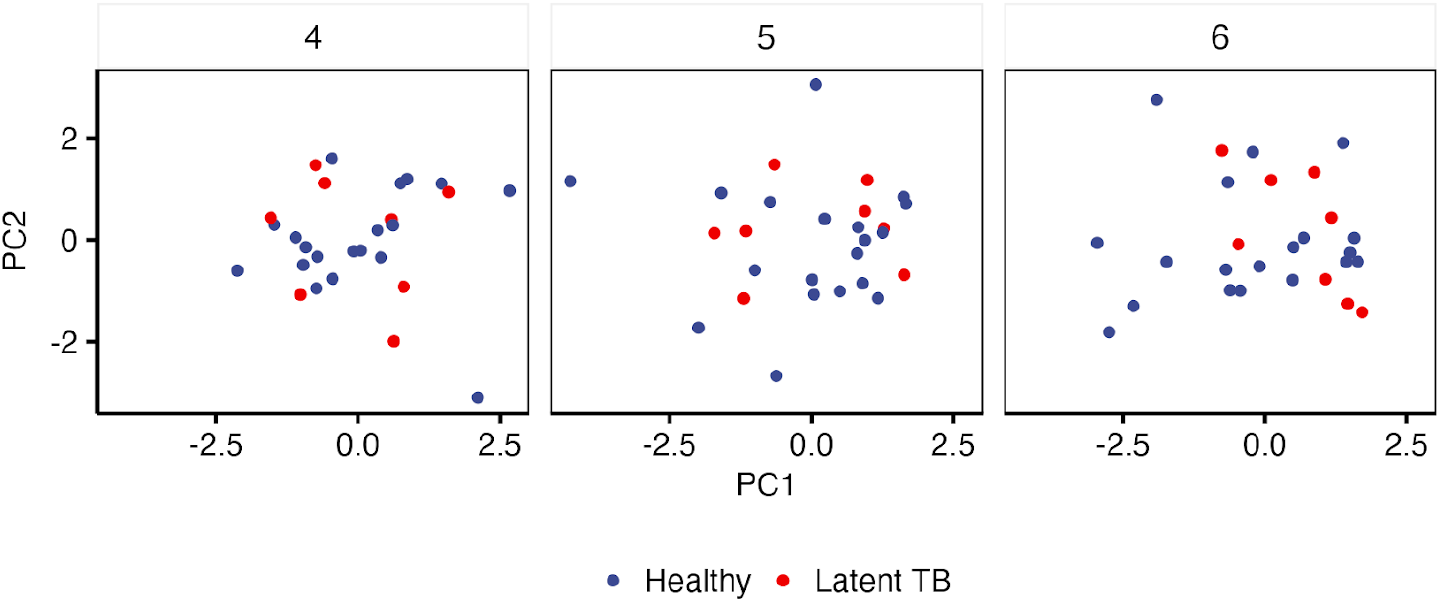
Principal component analysis of healthy and latent TB utilizing the derived biosignatures. X-axis shows the first component and Y-axis shows the second component.

**Supplementary Figure 5.**
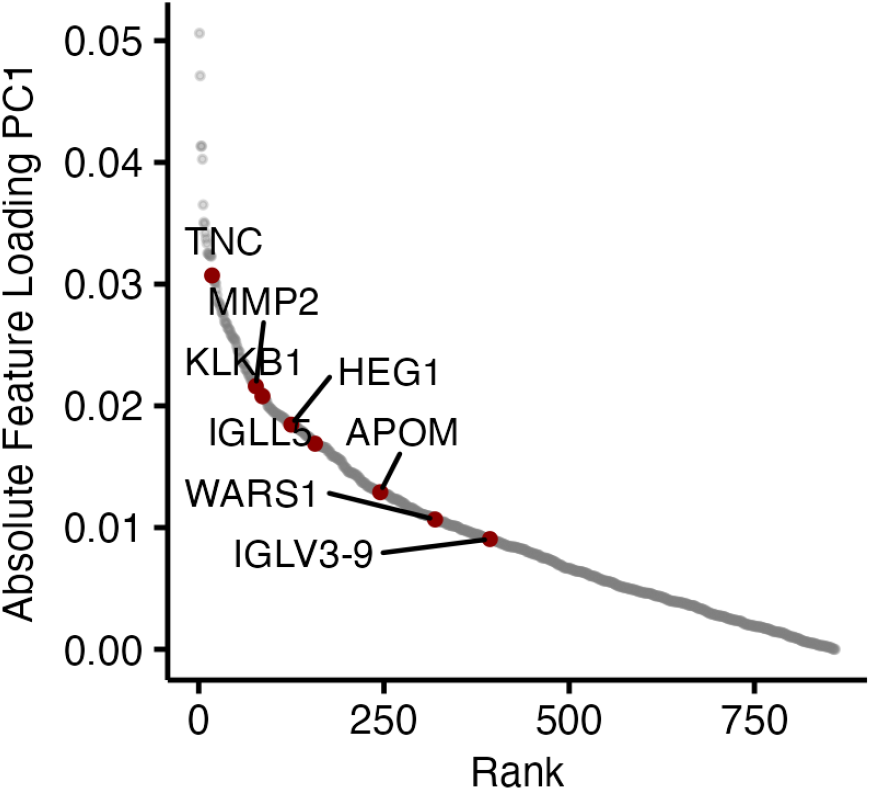
Rank Plot showing the contribution of each gene to the separation in the first component between confirmed + unconfirmed TB (all models). Y axis shows the feature loadings in absolute value and X axis displays the corresponding rank. Biosignature proteins are highlighted in red.

## Supplementary Tables

Supplementary Table S1. Clinical information table with clinical features and sample IDs.

Supplementary Table S2. Full proteomics output matrix of protein abundance by sample IDs.

Supplementary Table S3. Quantitative comparisons of Confirmed vs Unlikely TB.

Supplementary Table S4. Feature importance from lasso model.

## Material and methods

### Pediatric TB cohort

We analyzed plasma samples from children less than 15 years old evaluated for pulmonary TB as part of prospective diagnostic cohort studies in the Gambia, Peru, South Africa, and Uganda. Children were included if they had signs and symptoms of pulmonary TB, and excluded if they were already taking treatment for TB infection or disease for more than 72 hours. All children completed a standard TB evaluation, including clinical exam, chest X-ray, and respiratory sample collection for Xpert MTB/RIF molecular testing and mycobacterial culture. All children had follow up after 2-3 months, and were assessed for clinical response to any treatment. They were classified according to NIH consensus definitions as Confirmed, Unconfirmed, or Unlikely TB. Confirmed TB was defined as having microbiological evidence of TB disease by a positive Xpert MTB/RIF Ultra or mycobacterial culture positive for *M. tuberculosis*. Unconfirmed TB cases did not have microbiological evidence of TB, but had signs and symptoms of TB disease with other clinical signs or risk factors suggestive of TB including abnormal chest X-ray, known TB contact, and response to empiric anti-TB treatment. Unlikely TB cases were symptomatic, but did not have microbiological evidence of TB disease nor other signs or risk factors. In addition, asymptomatic healthy children from Uganda were enrolled, who had interferon-gamma release assay (IGRA) testing with Quantiferon-Gold (Qiagen, Hilden, Germany) testing for TB infection. Healthy controls were defined as asymptomatic and IGRA negative, while Latent TB infection cases were defined as asymptomatic with positive IGRA results. All caregivers completed a written informed consent, including for storage of samples for future studies, and children completed an assent as applicable. The studies were approved by the Mulago Hospital Ethics Research Committee, Gambian Government and MRC joint ethics committee, London School of Hygiene and Tropical Medicine, Institutional Ethics Committee for Research of National Institute of Health - Peru, University of Cape Town, and the University of California, San Francisco (UCSF) IRB.

### Sample Collection

Trained staff performed venipuncture and collected blood samples in all children at baseline and within 72 hours of any TB treatment. Blood samples were centrifuged and plasma samples aliquoted and placed in −80C freezers.

### Sample preparation for plasma proteomics

One uL of undepleted plasma was transferred in a 96 well plate with 200 uL of inactivation buffer (8M urea, 100 mM ammonium bicarbonate, 150 mM NaCl) and 0.75 uL/mL of RNAse (NEB) was added. The proteins were transferred to a 96 well filter plate and processed similarly to what we previously described^10^. Briefly, the plates were dried by centrifugation (1800 rpm at 25C for 30 minutes) and 50 uL of TUA buffer (8M urea, 20 mM ammonium bicarbonate, 5 mM TCEP) were added. Following incubation at RT on a shaker (500 rpm, 25 C), chloroacetamide (CAA) was added to 10 mM final concentration and the plates were incubated in the dark for 1 hr at room temperature. TCEP/CAA were removed by centrifugation and the plates were washed thrice with 200 uL of ddH20. Trypsin was added in a 1:50 ratio and the samples were digested overnight at 37°C on a shaker (800 rpm). Peptides were collected by centrifugation and the plate was washed once with 100 uL of ddH20. Resulting peptides were dried under vacuum and were resuspended at approximately 200 ng/uL. A representative pool of HIV positive and TB positive samples was further high-pH fractionated on a C18 tips following our previous work^20^.

### Data acquisition for abundance proteomics

Approx 200 ng per sample were analyzed on a Bruker TimsTOF Pro interfaced with a Ultimate 3000 UHPLC. Peptides were separated using a 15 cm PepSep column (Bruker, 150 cm length, 1.7 um beads) and sprayed into the Captive source kept at 1700 V and 200 C. The peptides were separated from 2 to 33% of buffer B (0.1% FA in ACN) for 26 minutes, then B was increased to 90% buffer B for 5 minutes, and then the column was re-equilibrated at 5% buffer B for 2 minutes, reaching a total gradient time of 33 min. The samples were acquired in DIA-PASEF mode using nine 32 m/z DIA-PASEF windows (500-966 mz) and ion mobility between 0.85 and 1.3 Vs/cm2.

### DDA-PASEF data analysis

Library files were searched using MSfragger^21^ within the FragPipe toolkit using the library generation workflow (‘DIA-Speclib-quant’) using a human FASTA (20408 entries). The generated library and our previous reported plasma library were merged using easypqp (https://github.com/grosenberger/easypqp).

### DIA-PASEF data analysis

All samples were searched with DIA-NN (v1.8)^22^ using a library-based strategy. MS1 and MS2 tolerances were set to 10 ppm. Protein grouping was performed based on the library ids and cross run-normalization was disabled. Following search, the global report file was filtered to <= 1% protein group Q-values (‘Lib.PG.Q.Value’) The peptide-level data was normalized using median-centering of the peptides identified in all samples.

Following normalization, the missing values were imputed utilizing an heuristic strategy based on their identification frequency to leverage the large number of samples analyzed in this study. The following rules were applied:

- Peptides identified in > 50% of the samples (at least 250 independent identifications) were imputed with the mean identification value,
- Peptides identified in < 50% but > 10% of the samples were imputed utilizing a random value extracted from a generated gaussian distribution with mu and sigma of the data downshifted 1.8 x sigma
- Peptides identified in < 10% of the samples were removed.

Following imputation, the peptide-level data was batch corrected using COMBAT^13^ to normalize any variation between the clinical sites, batches of sample preparation, or MS acquisition batches. We used as batches the various clinical sites, with added covariates of the MS acquisition and sample preparation batches (i.e the different plates). Peptides were rolled into proteins utilizing only proteotypic peptides and a topN strategy (max 3 proteotypic peptides per protein), using the mean intensity to represent a protein intensity.

### Machine learning for identification of a proteomic biosignature for childhood TB disease

Protein-level intensities after normalization across all clinical sites and HIV status for Confirmed TB (n=120) and Unlikely TB (n=211) were selected and z-scored. Confirmed TB and Unlikely TB cases were included given clear reference standards for TB and not TB. First, a random 75% of the data was selected for training a LASSO model using scikit-learn LASSOCv function (20 folds stratified by TB class, max_iter=10000, tol=0.0001). The feature importance was calculated and the proteins with non 0 coefficients were used for combinational analysis (n=67 proteins). In this analysis, we generated all possible combinations of features ranging from 1 (67 combinations) to 6 (n= 99795695 combinations) and trained a linear regression model based on the z-scored abundance for each specific combination. Models for every N were ranked based on the sensitivity achieved at 90% specificity (on our 25% test split) and the top scoring models for every N were kept for subsequent analysis. Confidence intervals were calculated using the Clopper-Pearson (exact binomial) method. We then applied models achieving the required WHO TPP (4, 5, and 6 protein models) to the Unconfirmed TB cases to determine what proportion could be diagnosed using this model.

